# Impact of anemia on length of hospital stay among patients hospitalized for severe malaria in Brazzaville, Republic of the Congo

**DOI:** 10.64898/2026.10.14.26365597

**Authors:** Martin Herbas Ekat, Amona Médard, Margaux Andzi Barré, Princesse Bendentt Lebaho, Gilius Axel Aloumba, Tatia Adoua Doukaga

## Abstract

**Background:** Severe malaria remains a major cause of hospitalization in malaria-endemic sub-Saharan Africa, and anemia is among its most frequent severity criteria. Data on the impact of anemia on hospital length of stay (LOS) among adults with severe malaria remain limited in Central Africa. We assessed the impact of anemia on LOS among adults hospitalized for severe malaria in Brazzaville, Republic of Congo.

**Methods:** We conducted a cross-sectional, descriptive, and analytic study of medical records from adult patients (≥18 years) diagnosed with severe malaria (WHO criteria) and admitted to four hospitals in Brazzaville between January 2020 and December 2024. We dichotomized LOS as short (<5 days) or prolonged (≥5 days). Multivariable logistic regression, using a backward stepwise procedure, identified factors independently associated with prolonged LOS; isolated hemoglobinuria served as the reference severity category.

**Results:** Among 523 patients hospitalized for severe malaria, 296 met the inclusion criteria. Median age was 25.5 years (IQR 20–40); 51.7% were female, and 82.1% had a low socioeconomic status. Median hemoglobin was 10.55 g/dL, and median LOS was 4 days (IQR 3–5). In multivariable analysis, prolonged LOS was independently associated with delay to consultation (aOR 1.25/day, 95% CI 1.09–1.44), isolated anemia (aOR 7.07, 1.23–40.55), convulsions (aOR 5.86, 1.81–18.97), prostration (aOR 4.42, 1.21–16.2), and impaired consciousness (aOR 4.29, 1.17–15.7). The strongest associations involved combined severity criteria: anemia with prostration (aOR 8.58), anemia with jaundice or hemoglobinuria (aOR 16.21), and impaired consciousness with anemia (aOR 33.31).

**Conclusions:** Anemia is a major, potentially modifiable determinant of prolonged hospitalization in adults with severe malaria, with a markedly amplified effect when combined with neurologic impairment. Early anemia screening, improved access to safe blood transfusion, and reduced delays to care could shorten hospital stays and ease the burden on health systems in this setting.

## Introduction

Malaria remains, after more than two decades of control efforts, the leading parasitic disease worldwide, responsible for substantial morbidity and mortality. According to the latest WHO World Malaria Report, an estimated 282 million cases and 610 000 deaths occurred in 2024, approximately 95% of both in the WHO African Region[1]. Severe malaria, defined by the presence of asexual Plasmodium forms together with at least one clinical or biological severity criterion, accounts for an estimated 15–18.8% of malaria cases in hospital-based series[2,3], with high case fatality in the absence of prompt, appropriate management, as illustrated by recent reports of death from severe malaria diagnosed late[4].

Among the WHO severity criteria, anaemia holds a particular place: it is one of the most frequent complications of severe malaria and a major contributor to its associated morbidity[3]. It occurs at broadly similar frequencies in Plasmodium falciparum and Plasmodium vivax infection, even though the latter is generally associated with lower parasitaemia, underscoring the multifactorial nature of malarial anaemia, which combines haemolysis, splenic sequestration, and dyserythropoiesis[5].

In the Republic of Congo, Plasmodium falciparum is by far the predominant species, accounting for the large majority of documented malaria infections in Brazzaville[6,7]. Malaria remains a major cause of hospitalisation in the country[8], and anaemia, particularly in children, is among the most frequently reported severity criteria, as shown in several studies from the Centre Hospitalier Universitaire de Brazzaville[9,10]. Although severe malarial anaemia is well documented clinically and pathophysiologically, its specific effect on length of hospital stay, particularly in adults, remains poorly studied in Central Africa. Length of stay is a simple, directly measurable indicator of the burden severe malaria places on health systems, which are often constrained in staffing[11], hospital beds, and labile blood products in this subregion[12].

It is in this context that we undertook this study, which aimed to assess the impact of anaemia on length of hospital stay among adult patients hospitalised for severe malaria in four health facilities in Brazzaville, Republic of Congo.

## Patients and Methods

### Study setting

The study was conducted in the infectious diseases departments of the Centre Hospitalier Universitaire de Brazzaville, the Makélékélé and Talangaï referral hospitals, and the internal medicine department of the Hôpital de Base de Bacongo. These facilities serve densely populated urban areas and represent different levels of the health-care pyramid.

### Study design

This was a cross-sectional, descriptive, and analytic study of medical records of patients admitted to the above-mentioned hospitals in Brazzaville between January 2020 and December 2024.

### Study population

The study included all male and female patients aged 18 years or older diagnosed with severe malaria at the selected hospitals during the study period. Only records documenting anaemia and length of hospital stay—the variables required for this study—were retained.

Inclusion criteria were:

- records with haemoglobin and haematocrit values;
- records documenting length of hospital stay, admission date, and discharge date;
- records with demographic data (age, sex, and socioeconomic status).

Exclusion criteria were:

- absence of parasitological confirmation of malaria (thick blood film and blood smear) in the patient’s record;
- confirmed malaria with a concurrent condition that could itself cause anaemia (eg, tuberculosis).

### Sample size

This was not an a priori calculated sample but an exhaustive census of all available severe malaria records during the study period. Of 523 patients hospitalised for severe malaria, 296 records meeting the inclusion criteria were reviewed and retained for analysis.

### Laboratory investigations

All participating hospitals had a laboratory able to perform thick blood film/blood smear examination and haemoglobin measurement.

### Parasite detection

In line with National Malaria Control Programme recommendations, thick blood films were prepared from capillary or EDTA-anticoagulated venous blood. Capillary blood was taken from the left ring finger. A large drop of blood, labelled with the patient’s details, was placed at the centre of the slide for the thick film, and a second drop near one end for the thin smear, following WHO methodology[13]. Thick films were stained for 15 min with 10% Giemsa solution diluted in mineral water; smears were fixed in methanol and stained with May-Grünwald-Giemsa for 15 min. Microscopy was done at 1000× magnification (oil-immersion objective). The thick film allowed detection and counting of Plasmodium parasites, while the smear allowed species identification. Parasite density was estimated per µL of blood, referenced to 6000 leucocytes.

### Haemoglobin measurement

Haemoglobin was measured on a haematology analyser from blood samples.

### Case definition

Severe malaria was defined as the presence of asexual Plasmodium falciparum forms in peripheral blood together with at least one WHO-defined severity criterion[14]. Severity criteria for each patient were established from clinical and paraclinical findings recorded at diagnosis, and included:

- Impaired consciousness: Glasgow Coma Scale score <11 in adults.
- Prostration: generalised weakness preventing the patient from sitting, standing, or walking unassisted.
- Multiple convulsions: more than two episodes within 24 h.
- Acidosis: base deficit >8 mEq/L or, if unavailable, plasma bicarbonate <15 mmol/L or venous plasma lactate ≥5 mmol/L; severe acidosis manifests clinically as respiratory distress (rapid, deep, laboured breathing).
- Hypoglycaemia: blood or plasma glucose <2.2 mmol/L (<40 mg/dL).
- Severe malarial anaemia: haemoglobin <7 g/dL or haematocrit <20% in adults, with parasite density >10 000/µL.
- Renal impairment: plasma or serum creatinine >265 µmol/L (3 mg/dL) or blood urea >20 mmol/L.
- Jaundice: plasma or serum bilirubin >50 µmol/L (3 mg/dL) with parasite density >100 000/µL.
- Pulmonary oedema: confirmed radiologically, or oxygen saturation <92% on room air with respiratory rate >30/min, often with chest indrawing and crackles.
- Significant haemorrhage: recurrent or prolonged bleeding from the nose, gums, or venepuncture sites; haematemesis or melaena.
- Shock: compensated shock is defined by capillary refill time ≥3 s or a proximal-to-distal limb temperature gradient without hypotension; decompensated shock by systolic blood pressure <70 mmHg in children or <80 mmHg in adults, with signs of hypoperfusion (cold extremities or prolonged capillary refill).
- Haemoglobinuria: cola-coloured or dark urine.
- Hyperparasitaemia: P. falciparum parasite density >10% of red cells or >100 000 parasites/µL.

Length of hospital stay was defined as the number of days between admission and discharge, and was categorised as short (<5 days) or long (≥5 days) for analysis.

### Data collection procedure

This consisted of an exhaustive review of medical records of severe malaria admissions at the participating hospitals, from which records of adult patients meeting WHO diagnostic criteria for severe malaria were retained.

### Data collection tools

Data were collected using a pre-established survey form documenting sociodemographic characteristics, diagnostic criteria, and treatment and outcome characteristics.

### Statistical analysis

Data were entered in Excel® and transferred to Stata 12® (College Station, TX, USA) for analysis.

Patient characteristics at admission were summarised as mean (95% CI) for normally distributed continuous variables or median (IQR) otherwise, and as counts and percentages for categorical variables. Variables were compared by length of stay (<5 days vs ≥5 days) using the χ² test or Fisher’s exact test for categorical variables, and the Mann-Whitney U test for continuous variables.

Logistic regression, with backward stepwise selection, was used to identify factors independently associated with prolonged hospitalisation (length of stay ≥5 days). Variables with p<0.20 on univariable analysis were retained for the multivariable model. Results are presented as odds ratios (OR) with 95% CIs. The proportional-odds assumption for each retained variable was assessed using the Schoenfeld residuals test.

Statistical significance was set at two-sided p<0.05 throughout.

### Ethics

This study was conducted in accordance with the Declaration of Helsinki. The Research Ethics Committee for Health Sciences (CERSSA) was consulted; because the study reviewed existing medical records, CERSSA determined that formal approval was not required, provided patient anonymity was maintained.

## Results

### Patient characteristics

Overall, 523 cases of severe malaria were hospitalised; 296 patient records were selected and included in the study.

Median age was 25.5 years (IQR:20–40); the 20–29-year age group was the most represented (109 cases, 36.8%). 153 patients (51.7%) were female; 243 (82.1%) had a low socioeconomic status. Mean time to consultation was 2.93 days (IQR:2.69–3.16); 88 patients (29.7%) had received treatment before hospitalisation. Clinically, median body temperature was 37.98°C (37.86–38.1), fever was present in 286 patients (96.6%), and coma and convulsions were present in 58 (19.6%) and 82 (27.7%), respectively. On biological assessment, median parasite density, haemoglobin, and haematocrit were 3000/µL (IQR:280–18 000), 10.55 g/dL (IQR:7.9–12.3), and 31.77% (IQR:22.55–37), respectively.

Artesunate was used to treat 278 patients (93.9%); 39 patients (13.2%) received a blood transfusion. One death was recorded. Further details are in Table 1 and Figure 1.

**Figure 1:**
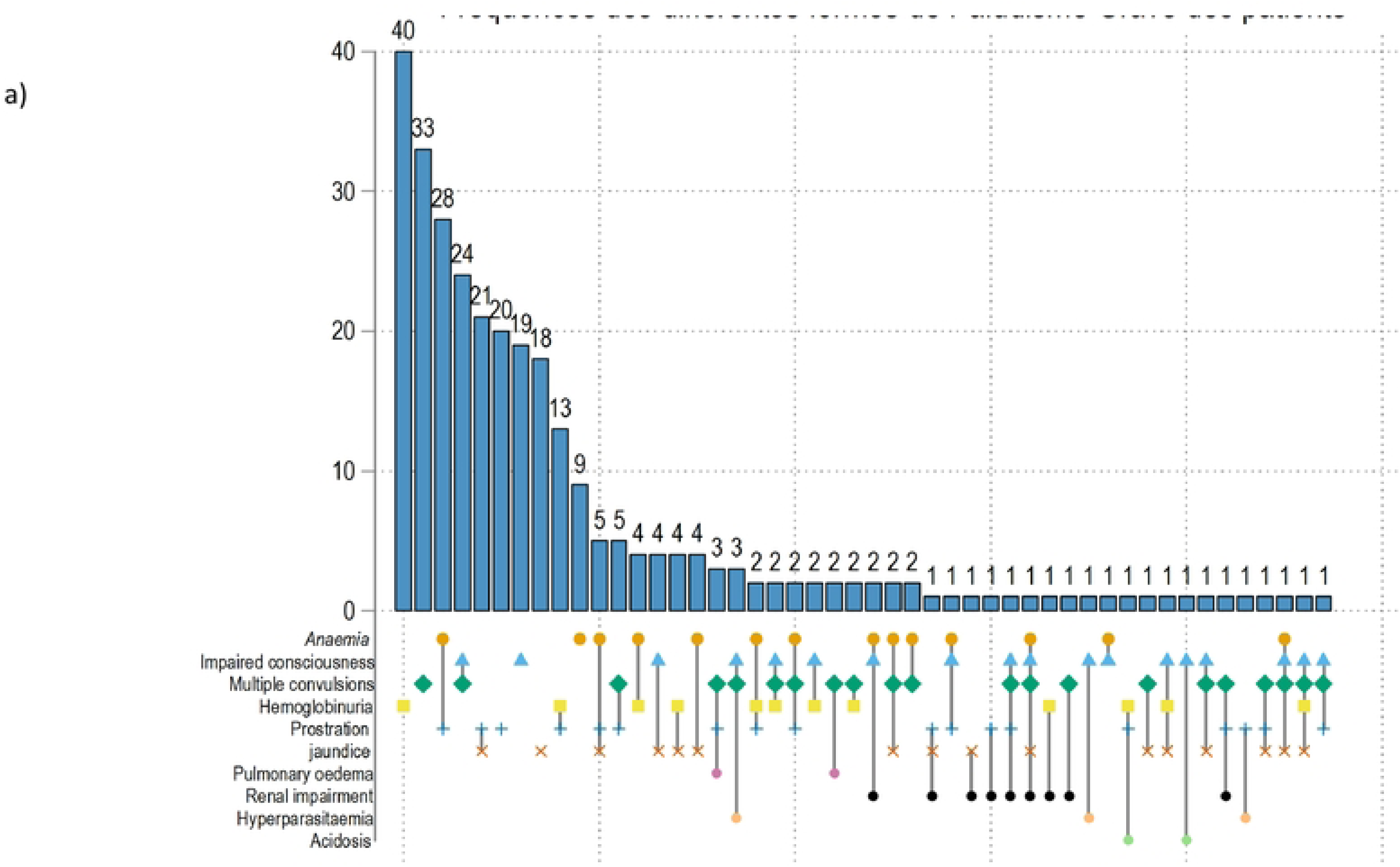
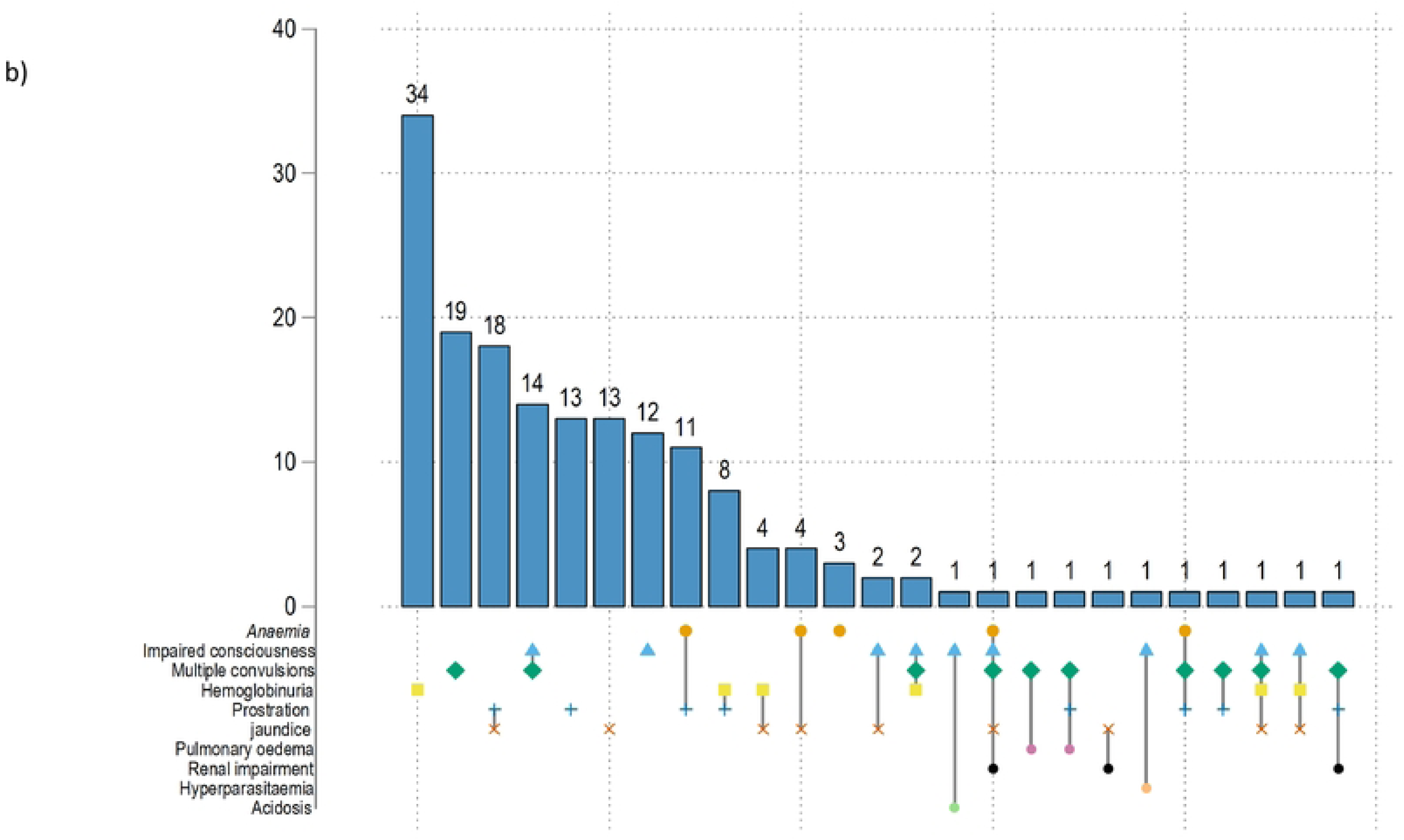
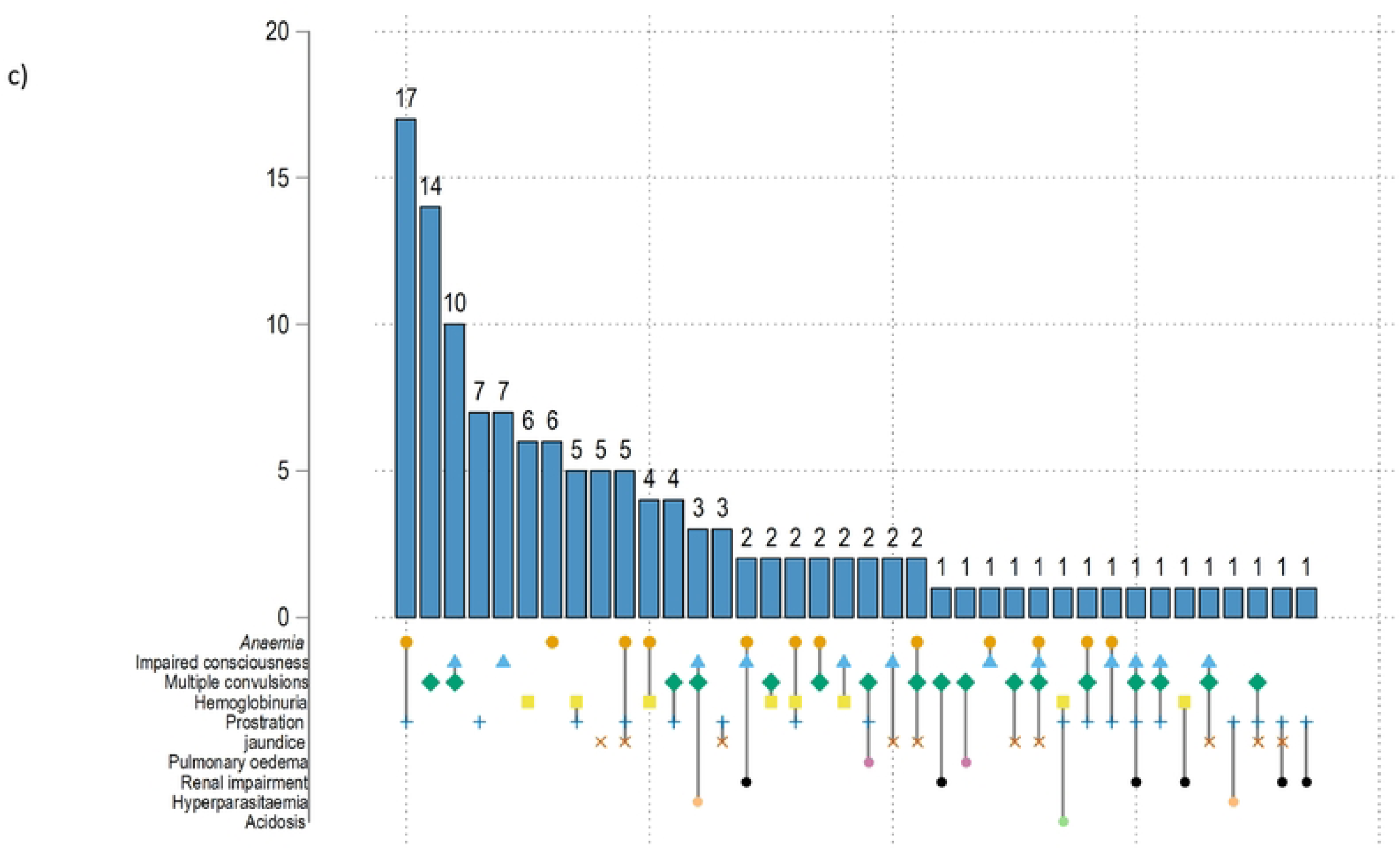
Frequency of severity criteria of severe malaria among patients hospitalised in the different hospitals of Brazzaville. (a) All patients. (b) Patients with short stay. (c) Patients with long stay.

**Table 1:** Characteristics of patients hospitalised for severe malaria in Brazzaville, between January 2020 and December, 2024.

| Variables | Total (N=296) | Short stay (1–4 days) (n=168) | Long stay (5 days or more) (n=128) | p value |
| --- | --- | --- | --- | --- |
| Length of hospital stay (days), median (IQR) | 4 (3 – 5) | 3 (3 – 4) | 6 (5 – 7) | $<0.0001$ |
| Age (years), median (IQR) | 25.5 (20 – 40) | 26 (20 – 36) | 25 (20 – 45.5) | 0.12 |
| Age group |  |  |  |  |
| < 20 years, n (%) | 70 (23.65) | 39 (23.21) | 31 (24.22) | 0.15 |
| 20 – 29 years, n (%) | 109 (36.82) | 61 (36.31) | 48 (37.5) |  |
| 30 – 39 years, n (%) | 42 (14.19) | 30 (17.86) | 12 (9.38) |  |
| 40 – 49 years, n (%) | 29 (9.80) | 17 (10.12) | 12 (9.38) |  |
| 50 – 59 years, n (%) | 21 (7.09) | 12 (7.14) | 9 (7.03) |  |
| >60 years, n (%) | 25 (8.45) | 9 (5.36) | 16 (12.5) |  |
| Sex |  |  |  |  |
| Female, n (%) | 153 (51.69) | 82 (48.81) | 71 (55.47) | 0.26 |
| Male, n (%) | 143 (48.31) | 86 (51.19) | 57 (44.53) |  |
| Socioeconomic status |  |  |  |  |
| Middle/high, n (%) | 53 (17.91) | 36 (21.43) | 17 (13.28) | 0.07 |
| Low, n (%) | 243 (82.09) | 132 (78.57) | 111 (86.72) |  |
| Time to consultation (days), mean (95% CI) | 2.93 (2.69 – 3.16) | 2.66 (2.38 – 2.93) | 3.29 (2.89 – 3.69) | 0.004 |
| Treatment received before hospitalisation |  |  |  |  |
| Yes, n (%) | 88 (29.73) | 45 (26.79) | 43 (33.59) | 0.3 |
| No, n (%) | 208 (70.27) | 123 (73.21) | 85 (66.41) |  |
| Type of treatment received before hospitalisation (**) |  |  |  |  |
| Antimalarials, n (%) | 78 (88.64) | 42 (93.34) | 36 (83.72) | NS |
| Antimalarial and antibiotic, n (%) | 2 (2.23) | 1 (2.22) | 1 (2.33) |  |
| Antibiotic, n (%) | 1 (1.14) | 1 (2.22) | 0 (0) |  |
| Herbal remedy, n (%) | 1 (1.14) | 0 (0) | 1 (2.33) |  |
| Not specified, n (%) | 6 (6.82) | 1 (2.22) | 5 (11.61) |  |
| **Clinical signs** |  |  |  |  |
| Glasgow Coma Scale score, mean (95% CI) | 14.09 (13.84 – 14.35) | 14.39 (14.1 – 14.69) | 13.7 (13.25 – 14.14) | 0.004 |
| Coma |  |  |  |  |
| Yes, n (%) | 58 (19.59) | 28 (16.67) | 30 (23.44) | 0.15 |
| No, n (%) | 238 (80.41) | 140 (83.33) | 98 (76.56) |  |
| Impaired consciousness |  |  |  |  |
| Yes, n (%) | 75 (25.34) | 36 (12.16) | 39 (13.18) | 0.08 |
| No, n (%) | 221 (74.66) | 132 (44.59) | 89 (30.07) |  |
| Convulsions (*) |  |  |  |  |
| Yes, n (%) | 82 (27.7) | 41 (24.4) | 41 (32.03) | 0.2 |
| No, n (%) | 214 (72.3) | 127 (75.6) | 87 (67.97) |  |
| Body temperature (°C), median (IQR) | 37.98 (37.86 – 38.1) | 37.99 (37.82 – 38.15) | 37.97 (37.77 – 38.17) | 0.6 |
| Fever |  |  |  |  |
| Yes, n (%) | 286 (96.62) | 162 (96.43) | 124 (96.88) | NS |
| No, n (%) | 10 (3.38) | 6 (3.57) | 4 (3.13) |  |
| Parasite density (/µL), median (IQR) | 3000 (280 – 18000) | 3200 (280 – 18000) | 2000 (400 – 17850) | 0.1 |
| Haemoglobin (g/dL), median (IQR) | 10.55 (7.9 – 12.3) | 11.1 (9.4 – 13) | 9.1 (6 – 11.4) | <0.0001 |
| Haemoglobin (++) |  |  |  |  |
| < 7 g/dL, n (%) | 58 (20.14) | 18 (10.91) | 40 (32.52) | <0.0001 |
| ≥ 7 g/dL, n (%) | 230 (79.86) | 147 (89.09) | 83 (67.48) |  |
| Haematocrit (%), median (IQR) | 31.77 (22.55 – 37) | 34 (28 – 39) | 27 (18.6 – 34) | <0.0001 |
| Haematocrit (+) |  |  |  |  |
| < 20%, n (%) | 61 (21.79) | 19 (11.88) | 42 (35) | <0.0001 |
| ≥ 20%, n (%) | 219 (78.21) | 141 (88.13) | 78 (65) |  |
| Serum creatinine (µmol/L), median (IQR) (##) | 88 (67.6 – 145.78) | 82.72 (67.6 – 119) | 102.2 (67.76 – 199) | 0.1 |
| Treatment received |  |  |  |  |
| Artesunate, n (%) | 278 (93.2) | 161 (95.83) | 117 (91.41) | 0.1 |
| Artemether, n (%) | 18 (6.08) | 7 (4.17) | 11 (8.59) |  |
| Blood transfusion |  |  |  |  |
| Yes, n (%) | 39 (13.18) | 9 (5.36) | 30 (23.44) | <0.0001 |
| No, n (%) | 257 (86.82) | 159 (94.64) | 98 (76.56) |  |
| Number of blood units received, mean (95% CI) (†) | 2.36 (2.04 – 2.68) | 2.22 (1.71 – 2.74) | 2.4 (2 – 2.8) | 0.3 |
| Death, n (%) | 1 (0.34) | 1 (0.6) | 0 (0) | NS |

Legend: IQR=interquartile range. Min=minimum. Max=maximum. \*=more than two seizures within 24 h. NS=not significant. \*\*=88 patients. \*\*\*=severe malaria criteria per WHO definitions. +=measured in 280 patients. ++=measured in 288 patients. ##=measured in 75 patients. †=among the 39 transfused patients.

Mean time to consultation was significantly longer among patients with prolonged stay (3.29 days [95%CI:2.89–3.69] vs 2.66 [95%CI:2.38–2.93], p=0.04); the mean of Glasgow Coma Scale score were lower among patients with prolonged stay (13,7 (95%CI: 13,25 – 14,14) vs 14,39 (95%CI: 14,1 – 14,69), p<0.004); and median haemoglobin and haematocrit were lower among patients with prolonged stay (9.1 g/dL [IQR:6–11.4] vs 11.1 [IQR:9.4–13], p<0.0001, and 27% [IQR:18.6–34] vs 34% [IQR: 28–39], p<0.0001, respectively) (Table 1).

### Severity criteria of severe malaria

The most common isolated severity criteria were haemoglobinuria (40 patients, 13.5%), convulsions (33, 11.2%), impaired consciousness (19, 6.4%), jaundice (18, 6.1%), prostration (20, 6.8%), and anaemia (9, 3.0%). The most frequent combinations were anaemia with prostration (28, 9.5%), impaired consciousness with convulsions (24, 8.1%), and jaundice with prostration (21, 7.1%).

Among patients with prolonged stay, the most frequent criteria were anaemia with prostration (17, 13.3%), convulsions (17, 13.3%), and impaired consciousness (10, 7.8%). Among patients with short stay, the most frequent were haemoglobinuria (34, 20.2%), convulsions (19, 11.3%), jaundice with prostration (18, 10.7%), impaired consciousness with convulsions (14, 8.3%), and prostration (13, 7.7%). Full details are in Table 2 and Figure 1.

**Table 2:** Severity criteria among patients hospitalised for severe malaria in Brazzaville, January, 2020, to December, 2024.

| Variables | Total (N=296) | Short stay (1–4 days) (n=168) | Long stay (5 days or more) (n=128) | p value |
| --- | --- | --- | --- | --- |
| Haemoglobinuria, n (%) | 40 (13.51) | 34 (20.24) | 6 (4.69) | <0.001 |
| Haemoglobinuria and prostration, n (%) | 13 (4.39) | 8 (4.76) | 5 (3.91) |  |
| Haemoglobinuria and renal impairment, n (%) | 1 (0.34) | 0 (0) | 1 (0.78) |  |
| Convulsions, n (%) | 33 (11.15) | 19 (11.31) | 14 (10.94) |  |
| Convulsions and prostration, n (%) | 4 (1.35) | 1 (0.60) | 3 (2.34) |  |
| Convulsions, prostration and renal impairment, n (%) | 1 (0.34) | 1 (0.60) | 0 (0) |  |
| Convulsions and pulmonary oedema, n (%) | 2 (0.68) | 1 (0.60) | 1 (0.78) |  |
| Convulsions and jaundice, n (%) | 1 (0.34) | 0 (0) | 1 (0.78) |  |
| Convulsions and haemoglobinuria, n (%) | 2 (0.68) | 0 (0) | 2 (1.56) |  |
| Convulsions and anaemia, n (%) | 2 (0.68) | 0 (0) | 2 (1.56) |  |
| Convulsions, anaemia and prostration, n (%) | 2 (0.68) | 1 (0.60) | 1 (0.78) |  |
| Jaundice, n (%) | 18 (6.08) | 13 (7.74) | 5 (3.91) |  |
| Jaundice and prostration, n (%) | 21 (7.09) | 18 (10.71) | 3 (2.34) |  |
| Jaundice, prostration and renal impairment, n (%) | 1 (0.34) | 0 (0) | 1 (0.78) |  |
| Jaundice and haemoglobinuria, n (%) | 4 (1.35) | 4 (2.38) | 0 (0) |  |
| Jaundice and renal impairment, n (%) | 1 (0.34) | 1 (0.60) | 0 (0) |  |
| Impaired consciousness, n (%) | 19 (6.42) | 12 (7.14) | 7 (5.47) |  |
| Impaired consciousness and prostration, n (%) | 1 (0.34) | 0 (0) | 1 (0.78) |  |
| Impaired consciousness and jaundice, n (%) | 4 (1.35) | 2 (1.19) | 2 (1.56) |  |
| Impaired consciousness, jaundice and prostration, n (%) | 1 (0.34) | 0 (0) | 1 (0.78) |  |
| Impaired consciousness, jaundice and haemoglobinuria, n (%) | 1 (0.34) | 1 (0.60) | 0 (0) |  |
| Impaired consciousness and renal impairment, n (%) | 1 (0.34) | 0 (0) | 1 (0.78) |  |
| Impaired consciousness and hyperparasitaemia, n (%) | 1 (0.34) | 1 (0.60) | 0 (0) |  |
| Impaired consciousness and haemoglobinuria, n (%) | 2 (0.68) | 0 (0) | 2 (1.56) |  |
| Impaired consciousness and anaemia, n (%) | 1 (0.34) | 0 (0) | 1 (0.78) |  |
| Impaired consciousness, anaemia and prostration, n (%) | 1 (0.34) | 0 (0) | 1 (0.78) |  |
| Impaired consciousness, anaemia and renal impairment, n (%) | 2 (0.68) | 0 (0) | 2 (1.56) |  |
| Impaired consciousness, anaemia and jaundice, n (%) | 2 (0.68) | 0 (0) | 2 (1.56) |  |
| Impaired consciousness and acidosis, n (%) | 1 (0.34) | 1 (0.60) | 0 (0) |  |
| Impaired consciousness and convulsions, n (%) | 24 (8.11) | 14 (8.33) | 10 (7.81) |  |
| Impaired consciousness, convulsions and prostration, n (%) | 1 (0.34) | 0 (0) | 1 (0.78) |  |
| Impaired consciousness, convulsions and jaundice, n (%) | 1 (0.34) | 0 (0) | 1 (0.78) |  |
| Impaired consciousness, convulsions, jaundice and haemoglobinuria, n (%) | 1 (0.34) | 1 (0.60) | 0 (0) |  |
| Impaired consciousness, convulsions and hyperparasitaemia, n (%) | 3 (1.01) | 0 (0) | 3 (2.34) |  |
| Impaired consciousness, convulsions and haemoglobinuria, n (%) | 2 (0.68) | 2 (1.19) | 0 (0) |  |
| Impaired consciousness, convulsions, prostration and renal impairment, n (%) | 1 (0.34) | 0 (0) | 1 (0.78) |  |
| Impaired consciousness, convulsions, anaemia and jaundice, n (%) | 1 (0.34) | 0 (0) | 1 (0.78) |  |
| Impaired consciousness, convulsions, anaemia and jaundice, n (%) | 1 (0.34) | 1 (0.60) | 0 (0) |  |
| Impaired consciousness, prostration and pulmonary oedema, n (%) | 3 (1.01) | 1 (0.60) | 2 (1.56) |  |
| Anaemia, n (%) | 9 (3.04) | 3 (1.79) | 6 (4.69) |  |
| Anaemia and prostration, n (%) | 28 (9.46) | 11 (6.55) | 17 (13.28) |  |
| Anaemia and jaundice, n (%) | 4 (1.35) | 4 (2.38) | 0 (0) |  |
| Anaemia, jaundice and prostration, n (%) | 5 (1.69) | 0 (0) | 5 (3.91) |  |
| Anaemia and haemoglobinuria, n (%) | 4 (1.35) | 0 (0) | 4 (3.13) |  |
| Anaemia, haemoglobinuria and prostration, n (%) | 2 (0.68) | 0 (0) | 2 (1.56) |  |
| Prostration, n (%) | 20 (6.76) | 13 (7.74) | 7 (5.47) |  |
| Prostration and hyperparasitaemia, n (%) | 1 (0.34) | 0 (0) | 1 (0.78) |  |
| Prostration and renal impairment, n (%) | 1 (0.34) | 0 (0) | 1 (0.78) |  |
| Prostration, haemoglobinuria and acidosis, n (%) | 1 (0.34) | 0 (0) | 1 (0.78) |  |

### Factors associated with prolonged hospital stay

On univariable analysis, factors associated with prolonged stay included age >60 years (OR:2.34, 95% CI: 0.87–5.74, p=0.09), age 30–39 years (OR:0.50, 95% CI: 0.22–1.14, p=0.1), and time to consultation per day (OR:1.17, 95% CI: 1.04–1.31, p=0.01). Among severity criteria, prolonged stay was associated with impaired consciousness (OR: 3.92, 95% CI: 1.16–13.22, p=0.03), convulsions (OR: 4.47, 95% CI: 1.49–13.45, p=0.008), anaemia (OR:11.33, 95% CI: 2.21–58.15, p=0.004), and prostration (OR:3.92, 95% CI: 1.16–13.22, p=0.03). Combined criteria showed stronger associations: anaemia with prostration (OR: 8.76, 95% CI: 2.77–27.73, p<0.0001), impaired consciousness with convulsions (OR:5.26, 95% CI: 1.67–16.62, p=0.005), impaired consciousness and/or convulsions with anaemia (OR:28.33, 95% CI: 4.93–162.84, p<0.0001), impaired consciousness, convulsions, and haemoglobinuria (OR:11.33, 95% CI: 1.68–76.26, p=0.01), impaired consciousness and/or convulsions with hyperparasitaemia (OR:22.67, 95% CI: 2.15–239.32, p=0.009), and anaemia with jaundice or haemoglobinuria (OR:15.58, 95% CI: 3.71–65.53, p<0.0001).

On multivariable analysis, independent predictors of prolonged stay were time to consultation per day (aOR:1.25, 95% CI: 1.09–1.44, p=0.002), impaired consciousness (aOR: 4.29, 95% CI: 1.17–15.7, p=0.03), convulsions (aOR:5.86, 95% CI: 1.81–18.97, p=0.003), anaemia (aOR:7.07, 95% CI: 1.23–40.55, p=0.03), and prostration (aOR:4.42, 95% CI: 1.21–16.2, p=0.03). Combined criteria showed the strongest associations: anaemia with prostration (aOR:8.58, 95% CI: 2.51–29.4, p=0.001), impaired consciousness with convulsions (aOR:5.39, 95% CI: 1.59–18.27, p=0.007), impaired consciousness and/or convulsions with anaemia (aOR:33.31, 95% CI:5.47–202.73, p<0.0001), impaired consciousness, convulsions, and haemoglobinuria (aOR:17.02, 95% CI: 2.26–128.2, p=0.006), impaired consciousness and/or convulsions with hyperparasitaemia (aOR:35.94, 95% CI: 2.86–451.36, p=0.006), and anaemia with jaundice or haemoglobinuria (aOR:16.21, 95% CI: 3.61–72.8, p<0.0001). Full details are in Table 3.

**Table 3:** Univariable and multivariable analysis of factors associated with length of hospital stay ≥5 days among patients hospitalised for severe malaria in Brazzaville between January 2020 and December, 2024.

| Variables | OR (IC95%) | p-value | aOR (IC95%) | p-value |
| --- | --- | --- | --- | --- |
| **Age group** |  |  |  |  |
| <20 years | Ref |  | Ref |  |
| 20–29 years | 0.99 (0.54 – 1.81) | 0.97 | 1.09 (0.54 – 2.22) | 0.8 |
| 30–39 years | 0.5 (0.22 – 1.14) | 0.1 | 0.68 (0.26 – 1.77) | 0.4 |
| 40–49 years | 0.89 (0.37 – 2.13) | 0.79 | 1.29 (0.47 – 3.59) | 0.6 |
| 50–59 years | 0.94 (0.35 – 2.53) | 0.91 | 0.82 (0.26 – 2.6) | 0.7 |
| >60 years | 2.34 (0.87 – 5.74) | 0.09 | 2.61 (0.85 – 8) | 0.09 |
| **Sex** |  |  |  |  |
| Male | Ref |  |  |  |
| Female | 1.31 (0.82 – 2.07) | 0.26 | 1.39 (0.79 – 2.45) | 0.3 |
| **Socioeconomic status** |  |  |  |  |
| Middle/high | Ref |  |  |  |
| Low | 1.78 (0.95 – 3.34) | 0.07 | 1.56 (0.72 – 3.39) | 0.3 |
| Time to consultation, per 1-day increase | 1.17 (1.04 – 1.31) | 0.01 | 1.25 (1.09 – 1.44) | 0.002 |
| **Treatment received before hospitalisation** |  |  |  |  |
| No | Ref |  |  |  |
| Yes | 1.38 (0.84 – 2.28) | 0.21 |  |  |
| **Clinical signs** |  |  |  |  |
| Body temperature, per 1°C increase | 0.99 (0.79 – 1.24) | 0.9 |  |  |
| **Fever** |  |  |  |  |
| No | Ref |  |  |  |
| Yes | 1.15 (0.32 – 4.16) | 0.8 |  |  |
| Parasite density, per 100 additional parasites | 1 (0.99 – 1) | 0.24 |  |  |
| **Severity criteria** |  |  |  |  |
| Haemoglobinuria<sup>†</sup>, n (%) | Ref |  | Ref |  |
| Haemoglobinuria and prostration<sup>#</sup>, n (%) | 4.25 (1.08 – 16.71) | 0.04 | 3.76 (0.9 – 15.65) | 0.07 |
| Impaired consciousness<sup>††</sup>, n (%) | 3.92 (1.16 – 13.22) | 0.03 | 4.29 (1.17 – 15.7) | 0.03 |
| Impaired consciousness and jaundice<sup>××</sup>, n (%) | 4.25 (0.75 – 23.98) | 0.1 | 5.21 (0.9 – 30.16) | 0.07 |
| Impaired consciousness and anaemia<sup>##</sup>, n (%) | 28.33 (4.93 – 162.84) | <0.0001 | 33.31 (5.47 – 202.73) | <0.0001 |
| Convulsions<sup>†††</sup>, n (%) | 4.47 (1.49 – 13.45) | 0.008 | 5.86 (1.81 – 18.97) | 0.003 |
| Convulsions and prostration<sup>×</sup>, n (%) | 8.5 (1.16 – 62.09) | 0.04 | 7.7 (0.87 – 68.39) | 0.07 |
| Convulsions and pulmonary oedema, n (%) | 5.67 (0.31 – 103.45) | 0.2 | 6.94 (0.33 – 147.14) | 0.2 |
| Impaired consciousness and convulsions<sup>*</sup>, n (%) | 5.26 (1.67 – 16.62) | 0.005 | 5.39 (1.59 – 18.27) | 0.007 |
| Impaired consciousness, convulsions and haemoglobinuria#, n (%) | 11.33 (1.68 – 76.26) | 0.01 | 17.02 (2.26 – 128.2) | 0.006 |
| Impaired consciousness, prostration and pulmonary oedema, n (%) | 11.33 (0.88 – 145.52) | 0.06 | 10.93 (0.73 – 164.82) | 0.08 |
| Jaundice**, n (%) | 1.57 (0.42 – 5.88) | 0.5 | 1.45 (0.37 – 5.73) | 0.6 |
| Prostration***, n (%) | 3.92 (1.16 – 13.22) | 0.03 | 4.42 (1.21 – 16.2) | 0.03 |
| Anaemia, n (%) | 11.33 (2.21 – 58.15) | 0.004 | 7.07 (1.23 – 40.55) | 0.03 |
| hyperparasitaemia***, n (%) | 22.67 (2.15 – 239.32) | 0.009 | 35.94 (2.86 – 451.36) | 0.006 |
| Jaundice and prostration, n (%) | 1.26 (0.31 – 5.05) | 0.8 | 1.33 (0.31 – 5.64) | 0.7 |
| Anaemia and prostration, n (%) | 8.76 (2.77 – 27.73) | <0.0001 | 8.58 (2.51 – 29.4) | 0.001 |
| Anaemia and jaundice ; anaemia haemoglobinuria, n (%) | 15.58 (3.71 – 65.53) | <0.0001 | 16.21 (3.61 – 72.8) | <0.001 |

Legend: †=plus one case with haemoglobinuria and renal impairment. ††=plus one case each with impaired consciousness and prostration, impaired consciousness and renal impairment, and impaired consciousness and acidosis. †††=plus one case with convulsions and jaundice. \*=plus one case each with impaired consciousness, convulsions and prostration; impaired consciousness, convulsions and jaundice; and impaired consciousness, convulsions, prostration and renal impairment. \*\*=plus four cases with jaundice and haemoglobinuria, and one with jaundice and renal impairment. \*\*\*=plus one case with prostration and renal impairment, and one with prostration, haemoglobinuria and acidosis. ×=plus one case with convulsions, prostration and renal impairment. ××=plus one case each with impaired consciousness, jaundice and prostration; impaired consciousness, jaundice and haemoglobinuria; and impaired consciousness, convulsions, jaundice and haemoglobinuria. ×××=plus three cases with impaired consciousness and convulsions, one with impaired consciousness, and one with hyperparasitaemia. #=plus one case with haemoglobinuria and renal impairment. ##=two cases with convulsions and anaemia, two with impaired consciousness, convulsions and anaemia, two with convulsions, anaemia and prostration, and one with impaired consciousness, anaemia and prostration.

## Discussion

This cross-sectional, descriptive, and analytic study reviewed records of 296 patients hospitalised for severe malaria in four health facilities in Brazzaville between January 2020 and December 2024 to assess the impact of anaemia on length of hospital stay. The median length of stay was 4 days (IQR: 3–5). On multivariable analysis, with isolated haemoglobinuria as the reference category, prolonged stay (≥5 days) was independently associated with time to consultation (aOR:1.25 per day, 95% CI: 1.09–1.44, p=0.002) and with each severity criterion individually: anaemia (aOR:7.07,95% CI:1.23–40.55, p=0.03), convulsions (aOR:5.86, 95% CI:1.81–18.97, p=0.003), prostration (aOR:4.42, 95% CI: 1.21–16.2, p=0.03), and impaired consciousness (aOR:4.29, 95% CI: 1.17–15.7, p=0.03). Notably, combined severity criteria carried the highest adjusted odds ratios in the study: anaemia with prostration (aOR:8.58, 95% CI: 2.51–29.4, p=0.001), anaemia with jaundice or haemoglobinuria (aOR:16.21, 95% CI: 3.61–72.8, p<0.001), and, most strikingly, impaired consciousness with anaemia (aOR: 33.31, 95% CI: 5.47–202.73, p<0.0001). These findings confirm that anaemia is a major determinant of length of stay in adult severe malaria, and further suggest a cumulative, possibly synergistic, effect when combined with neurological impairment.

### Choice of reference category

Unlike our initial analysis, which grouped neurological severity criteria into a single category, this analysis distinguishes each severity criterion individually and in its main combinations, using isolated haemoglobinuria as the reference. This choice reflects haemoglobinuria’s clinical course in our series: it was the most frequent isolated criterion among patients with short stay (34 patients, 20.2%), consistent with acute haemolysis rapidly controlled by effective antimalarial treatment with little lasting systemic effect. Using this subgroup as reference allowed us to quantify, for each criterion or combination, the excess risk of prolonged stay relative to a relatively more favourable form of severe malaria, sharpening the clinical interpretation of the prognostic weight of WHO severity criteria on length of stay, which is clearly not uniform across criteria.

### Anaemia and length of hospital stay

Anaemia was central to our findings: median haemoglobin was significantly lower among patients with prolonged stay (9.1 g/dL vs 11.1 g/dL, p<0.0001), as was median haematocrit (27% vs 34%, p<0.0001). Alone, anaemia increased the odds of prolonged stay more than seven-fold relative to isolated haemoglobinuria. This association echoes recent hospital-based studies from endemic settings: in Madagascar, blood transfusion and high parasite density—both closely linked to anaemia severity—were among the factors most strongly associated with longer stay[11]; in Germany, median length of stay for imported malaria rose from 3 days overall to 7 days for severe cases, with transfusion or organ support prolonging hospitalisation[15]; and similar findings have been reported in Uganda and Kenya, where markers of haematological severity and anaemia-related investigations or procedures delayed discharge[16,17].

Pathophysiologically, severe malarial anaemia results from acute haemolysis of parasitised and non-parasitised red cells, splenic sequestration, and marrow dyserythropoiesis driven by systemic inflammation, as shown histologically in infected patients[18]. Unlike fever or parasitaemia, which resolve rapidly with effective treatment, haemoglobin recovery is slower, requiring either gradual marrow recovery or transfusion—with the delays in obtaining labile blood products that this entails in our setting. In our series, 13.2% of patients were transfused, rising to 23.4% among those with prolonged stay versus 5.4% with short stay (p<0.0001). A Cochrane review of blood transfusion for malarial anaemia reported that, in one controlled trial, early transfusion reduced length of stay compared with conservative management, while noting the risk of transfusion-related adverse events[19]. These findings align with the TRACT trials in Ugandan and Malawian children, which clarified optimal transfusion volume and timing in severe malarial anaemia while identifying excess mortality with larger transfusion volumes in febrile children[20,21]. Transfusion need thus appears to be both a marker of anaemia severity and an organisational determinant of length of stay, particularly where blood product availability is limited.

These findings are consistent with paediatric data from Brazzaville: in a study of children hospitalised for severe malaria at the Centre Hospitalier Universitaire de Brazzaville, anaemia was already the most frequent severity criterion, ahead of repeated convulsions and coma[9], and an earlier Congolese study specifically addressing anaemic severe malaria in children under five found a high prevalence of this clinical form, associated with low family socioeconomic status[10]. The convergence of these paediatric observations with our adult findings suggests that severe malarial anaemia remains, across all ages, a major and recurring component of the hospital burden of severe malaria in Brazzaville.

### A marked cumulative effect of anaemia and neurological impairment

The most striking finding of this analysis is the magnitude of the association observed when anaemia coexisted with impaired consciousness (aOR:33.31, 95% CI:5.47–202.73)—far exceeding either criterion alone (7.07 and 4.29, respectively). A comparable, though smaller, pattern was seen for anaemia with prostration (aOR 8.58) and for anaemia with jaundice or haemoglobinuria (aOR 16.21). This is consistent with a Colombian study in hospitalised adults with malaria, which found a significant interaction between admission haemoglobin and parasitaemia on the risk of multiple organ dysfunction syndrome, with risk markedly increased when severe anaemia and high parasitaemia coexisted but comparable to non-anaemic patients when parasitaemia was low[22]. More broadly, several studies have shown that the risk of multiple organ dysfunction and mortality in severe malaria rises disproportionately with the number of organs or systems involved, with one Indian intensive-care study reporting markedly higher mortality among patients with two or more organ systems affected than those with one[23]. Our findings extend this notion of cumulative burden to length of hospital stay: the combination of anaemia and neurological impairment does not appear to simply add two independent risks but may reflect an overall severity of parasitic insult—with concurrent erythrocytic and microvascular sequestration in the cerebral and marrow compartments—that further delays the clinical and biological recovery required for safe discharge.

### Impaired consciousness and convulsions

Alone, impaired consciousness and convulsions were each independently associated with prolonged stay (aOR 4.29 and 5.86, respectively), and their combination further increased this risk (aOR 5.39), as did their combination with haemoglobinuria (aOR 17.02) or hyperparasitaemia (aOR 35.94, though with a wide confidence interval given the small numbers involved). These findings are consistent with the literature: in the Apac study in Uganda, neurological manifestations were among the main determinants of prolonged stay in children with severe malaria[16] and the German study noted above found a similar increase in length of stay among patients with organ dysfunction—neurological, respiratory, or renal—all requiring close monitoring and gradual clinical recovery before safe discharge[15]. Recovery of consciousness after cerebral malaria is often delayed relative to parasite clearance because of cerebral microcirculatory injury associated with sequestration of parasitised red cells, accounting for the prolonged observation these patients require.

### Delay to care

In our study, each additional day of delay before the first consultation increased the odds of prolonged stay by 25% (aOR 1.25, 95% CI 1.09–1.44, p=0.002). This finding echoes guidance on the management of imported severe malaria, which emphasises that early diagnosis and prompt treatment remain essential determinants of prognosis[24,25]. In Morocco, El Mezouari and colleagues, studying cases of severe malaria admitted to intensive care, similarly found that diagnostic and therapeutic delay—often linked to late consultation or initial diagnostic uncertainty—remained a leading cause of clinical deterioration[26]. In our population, low socioeconomic status predominated (82.1% of patients), and nearly a third of patients had received treatment, usually antimalarial, before hospitalisation, reflecting frequent reliance on self-medication or first-level care before referral. These findings support strengthening community awareness of early warning signs of malaria and improving access to first-line health facilities, in line with WHO guidance on the management of severe malaria[14].

### Prostration

Prostration was also independently associated with prolonged stay, and its combination with anaemia increased this risk further. This is consistent with the Apac study in Uganda, where prostration, together with respiratory distress, was among the few factors independently associated with prolonged stay in children[16]. Prostration reflects a global deterioration in clinical status that often requires prolonged nutritional and fluid support before patients regain functional autonomy, which may explain both its independent contribution and its cumulative effect with anaemia on length of stay.

### Jaundice and haemoglobinuria: more rapidly resolving presentations

In contrast, isolated jaundice was not significantly associated with length of stay relative to isolated haemoglobinuria (aOR 1.45, p=0.6), nor was jaundice with prostration (aOR 1.33, p=0.7). This is consistent with our choice of isolated haemoglobinuria as reference: both manifestations reflect acute intravascular haemolysis and tend to resolve quickly with effective antimalarial treatment once parasite clearance is achieved, without requiring the prolonged care needed for recovery from severe anaemia or impaired consciousness. Only when anaemia coexisted with jaundice or haemoglobinuria did the risk of prolonged stay again become significantly elevated (aOR 16.21), confirming that the anaemic component of these combined haemolytic presentations, rather than haemolysis or jaundice per se, drives most of this risk.

### Strengths and limitations

This study has several strengths: its multicentre design across four facilities representing different levels of Brazzaville’s health-care pyramid, a relatively long inclusion period (5 years), and a detailed analysis of severity criteria, alone and in combination, that revealed the cumulative effect of anaemia with other severity criteria. It also has important limitations. Its retrospective design carries a risk of selection bias toward better-documented records, since only 296 of 523 patients hospitalised for severe malaria during the study period could be included. Fine stratification of combined severity criteria produced very small subgroups—sometimes fewer than five patients (eg, impaired consciousness with anaemia, or hyperparasitaemia)—yielding wide confidence intervals; the highest odds ratios should therefore be interpreted with caution, as signals to be confirmed rather than precise estimates. Some variables, such as creatinine or pre-admission treatment type, were available for only a subset of patients. Finally, the absence of post-discharge follow-up precludes assessment of anaemia’s impact on outcomes beyond discharge.

## Conclusion

In conclusion, anaemia emerges as one of the principal determinants of prolonged hospitalisation among adults with severe malaria in Brazzaville, whether alone or, more markedly, when combined with neurological impairment, prostration, or other haemolytic manifestations. These findings confirm the specific burden of severe malarial anaemia, already well documented in children in this city, and suggest that the hospital burden of severe malaria rises disproportionately with the accumulation of severity criteria. In light of these findings, early anaemia screening—particularly in patients with impaired consciousness or prostration—improved access to safe blood transfusion, and reduced delays to care represent priority levers for reducing length of stay and easing the burden severe malaria places on the Congolese health system.

## Data Availability

The datasets analyzed during the current study are available from the corresponding author upon reasonable request (deidentified participant data, data dictionary).

## Supporting information

**S1 Figure 1:** Frequency of severity criteria of severe malaria among patients hospitalised in the different hospitals of Brazzaville. (a) All patients. (b) Patients with short stay. (c) Patients with long stay.

## Acknowledgements

To the health workers in the departments visited across the four hospitals.

## Notes

### Competing Interest Statement

The authors have declared no competing interest.

### Author Declarations

The Research Ethics Committee for Health Sciences (CERSSA) was consulted because the study reviewed existing medical records, CERSSA determined that formal approval was not required, provided patient anonymity was maintained

